# Implementation of Tuberculosis Infection Control Practices in Tuberculosis Diagnostic and Treatment Health Facilities in Kampala District, Uganda, August 2015

**DOI:** 10.1101/2020.07.14.20119214

**Authors:** Didas Tugumisirize, Stavia Turyahabwe, Lilian Bulage, Stella Zawedde Muyanja, Robert Kaos Majwala, Michael Kakinda, Simon Muchuro, Abdunoor Nyombi, Andrew Nsawotebba, Denis Mujuni, Joel Kabugo, Simon Peter Katongole

**Author notes:** **Correspondence:** Didas Tugumisirize. **Email addresses** (Didas Tugumisirize), (Stavia Turyahabwe), (Lilian Bulage), (Stella Zawedde Muyanja), (Robert Kaos Majwala), (Simon Muchuro), (Michael Kakinda), (Abdunoor Nyombi), (Andrew Nsawotebba), (Dennis Mujuni), k (Joel Kabugo), (S. P. Katongole).

## Abstract

**Background:** Effective implementation of Tuberculosis infection control (TB IC) measures in health facilities delivering TB care services is very critical in controlling nosocomial transmission of TB infections among health workers, patients and their attendants. The aim of the study was to assess and document the implementation of TB IC practices in TB diagnostic and treatment health facilities in Kampala District, which accounts for 15-20% of the total TB burden in Uganda.

**Methods:** In August 2015, we conducted a cross-sectional study in 25 health facilities including 07 Public and 18 Private healthcare facilities in Kampala. We used a modified checklist adopted from the national manual for implementing TB control measures in health care facilities. We reviewed health facility records and where necessary observed TB IC practices to triangulate our findings. We conducted univariate analysis and generated proportions in order to describe the extent of implementation of TB IC measures.

**Results:** On average, 73% of both administrative and managerial, 65% environmental, and 56% personal protective TB IC measures were complied with at the health facilities visited. Private health facilities implemented 71% of both administrative and managerial TBIC measures compared to public health facilities (31%). Thirty Six percent of health facilities reported that they were regularly screening health care workers for TB. By Observation, 28% had TB IC guideline, 36% had TB IC plan, 12% had a designated area for sputum collection, 56% were regularly opening windows, 40% had fans installed in the waiting areas and/or consultation rooms and 24% had bio-safety cabinets fitted with UV light. In addition, 60% had N95 respirators but only 32% of the facilities reported that their health workers routinely wore them.

**Conclusion:** Implementation of WHO recommended TB IC measures in health facilities delivering TB care services in Kampala was sub optimal. Routine involvement of health facility management as well as increasing human resources for health is critical in implementing easy to do TBIC measures like triaging, patients’ educating on coughing etiquette and respiratory hygiene and daily window opening particularly in public health care settings where implementation of administrative TB IC measures is wanting

## Background

Effective Tuberculosis (TB) prevention and control measures are a critical part of the quality of health service delivery to achieve people-centered integrated universal health coverage. There has been accelerated efforts to control TB worldwide as evidenced by the recent global decline in the annual TB incidence rates [1]. In order to sustain these efforts towards ending TB, all high TB burden countries are expected to reach a 50% and 90% reduction in TB incidence rates by 2025 and 2035 respectively, compared with levels in 2015 [2]. However, these ambitious global incidence targets are most unlikely to be achieved by low and middle income countries due to huge resources required to systematically and extensively implement Tuberculosis infection control (TB IC) measures in health care settings and other congregate setting. In this particular study, Tuberculosis infection control was defined as a combination of managerial, administrative, environmental and personal protective measures aimed at minimizing the risk of TB transmission within health facilities delivering TB care services.

In Uganda, the national TB infection control efforts have been further curtailed by association of TB with HIV and the emergence of multi-drug resistant TB (MDR-TB) and extensively drug-resistant TB (XDR-TB). The country ranks 17^*^ among the first 30 high TB/HIV burdened countries in the World [3], with a TB/HIV co-infection rate of about 40% [4] and a high HIV prevalence of 7.3%. In addition, the prevalence of TB disease in Uganda is almost twice higher than had been previously estimated by World Health Organization, 253 compared to 161 per 100,000 population [5]. Similarly, the incidence of TB among health workers is estimated at 593 per 100,000 which is much higher than the general population rate of 200 per 100,000 [6]. Worse still, over 56% of all TB cases notified to the National TB and Leprosy program (NTLP) annually are bacteriologically confirmed incident TB cases [4]. This points to an ongoing high transmission of TB infections in health care settings and general populations respectively. The risk of transmission of tuberculosis in health care facilities from individuals with TB to other patients and to health professionals has been recognized for decades in many other low and middle income countries [7],[8].

In response to the increasing global public health concern of nosocomial TB infections, the WHO developed and disseminated TB IC guidelines in 2009 and further recommended all high TB burden countries to implement four levels of TB infection control (i.e. managerial, administrative, environmental and personal protective measures) in all congregate settings including health facilities [9]. Following this WHO directive, in 2011, Uganda’s Ministry of Health successfully adopted and disseminated this policy to all health facilities delivering TB care services [10]. These, if implemented in an integrated manner, have been shown to reduce and prevent the risk of transmission and exposure to tuberculosis [11, 12, 13]. However, the extent of implementation of such TBIC measures in health facilities delivering TB care services since the official launch of TBIC guidelines in the country is unknown. This post evaluative study is the first of its kind to assess the Implementation of TBIC practices in health facilities in Kampala, Uganda. Only one other similar study had been conducted prior to the release of Uganda TBIC guidelines, moreover, in semi-urban areas of Mukono and Wakiso districts in Uganda [14]. In addition to the WHO recommended TB IC measures, we also assessed the waste management practices in health facilities delivering TB care services. It is anticipated that the findings of this study may be used by health planners, institutional mangers of National TB programs to review their TB IC practices and design appropriate interventions to address the bottlenecks identified.

## Methods

### The study design and setting

A cross-sectional study was conducted in August 2015 in selected TB diagnostics and treatment Health Care Facilities in Kampala district. The study area was selected because it accounts for 15-20% of the total TB notifications in Uganda [15]. Further, TB disease is associated with overcrowding conditions and yet Kampala has the highest number of populations among all urban centers in the country, accounting for 25% of the national urban populations in the country [16]. Thus, health facilities based in such urban settings usually receives huge numbers of outpatients despite the low health worker patient ratio demonstrated resource limited health care settings, which in turn results into long waiting time and thus increased risk of TB transmission

### Sample Size and Sample Size Determination

Hyper geometric formula was used to determine the sample size of the study. A total of 49 TB diagnostic and treatment health care facilities were sampled to participate in this study.

### Sampling procedure

A multi stage sampling technique was employed in this study. Following the establishment of the required sample size (n=49), the first stage of sampling involved determining the desired number of health care facilities per division using “Probability proportional to size (PPS)” technique such that all the divisions in Kampala were represented equally in the study. Further, to minimize sampling bias and ensure that every DTU had an equal chance of participation, the next stage included selection of one health care facility at a time from each of the 6 divisions through a simple random sampling without replacement technique using paper ballots until the desired sample size per division is reached.

### Eligibility Criteria

All health facilities in Kampala district, which at the time of the study, were directly involved in diagnosis and treatment of TB as guided by Uganda’s NTLP database were included in the study. Respondents of the TBIC assessment checklist included health care facility in-charges, laboratory technicians, health facility TB focal persons and/or TB/HIV coordinators from the selected health facilities in Kampala District. Health facilities whose targeted respondents were absent during data collection period were excluded from the study

### Data Collection

In order to assess whether the recommended TBIC practices were being implemented in health care facilities, we modified and used a checklist adopted from a standardized tool for assessment of adherence to recommended TBIC from the manual for implementing TB control measures in health care facilities [17]. Respondents for this modified checklist were health care facility in-charges and in their absence, health facility TB focal persons and TB/HIV coordinators. This data collection tool sought information on the characteristics of the health facility (i.e. facility level and ownership) and TBIC measures available based on the recommended Uganda 2011 guidelines for TB Infection Control in health care facilities, congregate settings and households [10].

Administrative TBIC measures assessed included all initiatives taken by health care facility managers/administrators to curtail TB transmission within the health facility such as availability of TBIC guidelines, committee and plan, patient education and/or counselling about TB, availability of IEC materials, early diagnosis and prompt treatment of TB patients, screening, separation and prioritization of presumptive TB patients, presence of designated and well ventilated areas for sputum collection, health care facility assessments and infrastructure maintenance activities, TBIC trainings and regular screening of health workers for TB. On the other hand, environmental TBIC measures assessed included all those methods put in place not only to reduce the concentration of infectious agents in the air but also control the direction of flow of air in and out of a health facility room so as to prevent health workers, patients, attendants and visitors at the health facility from inhaling air contaminated with infectious TB agents. These included; Adequate natural ventilation (i.e. at the waiting areas, laboratory and clinical rooms), availability of mechanical air-moving equipment (e.g. use of window fans in clinical rooms), availability of bio-safety cabinets fitted with ultra-violet (UV) light in the laboratory and waste management practices at the health facility. Ultimately, we investigated the presence of personal protective gears such as N95 and whether they were persistently worn by health workers.

In order to have a more objective assessment, direct observations of TBIC measures were carried out using an observation checklist [14], [9]. Further review of health facility documents and records was carried out where necessary. Some of the documents reviewed included, minutes of previous CMEs conducted on TBIC, TB infection control plan to verify its components, TB contact tracing and treatment registers, TB laboratory registers and intensified TB case finding forms.

### Quality Control Procedures

The study tools were pretested and revised accordingly. Experienced research assistants were trained on the objectives of the study and how to administer the tools. Reliability of the study findings was ensured by triangulation involving use of observations checklist and health facility TBIC assessment checklist to collect health care facility survey data as well as review of health facility documents and records where necessary

### Data Management and Analysis

Prior to analysis, all the filled out health care facility (HCF) assessment checklists were coded for confidentiality purposes. We conducted univariate analysis and generated proportions in order to ascertain the extent of implementation of TBIC measures. In addition, using self-reported data, the number of TB diagnostics and treatment health care facilities implementing each TB Infection Control measure were stratified according to level of ownership.

## Results

Overall, 25 TB diagnostic and treatment health care facilities participated in this study. Of these, 28% (7/25) were public and 72% (18/25) privately owned. Among public health facilities, majority 57% (4/7) were HC IIIs, 29% (2/7) were hospitals and only 14% (1/7) were HC IVs. All the health care facilities were assessed using a TBIC checklist, an observation checklist and where necessary reviewed health care facility records. On average, 73% administrative and managerial, 65% environmental and 56% personal protective measures were complied with at the health facilities visited [Table 1]. About 71% of the administrative TB IC measures were complied with at private health facilities compared to 31% at Public health facilities [Table 2]. Notable discrepancies were observed between health facility survey data and observed data [Table 3]

**Table 1:** Extent of Implementation of TB IC Measures in Health Facilities delivering TB services in Kampala

| HEALTH FACILITY AND OWNERSHIP |  | LEVEL OF TB INFECTION CONTROL IMPLEMENTED |  |  |  |  |  |
| --- | --- | --- | --- | --- | --- | --- | --- |
|  |  | Administrative/Managerial (n = 18) |  | Environmental (n = 7) |  | Personal Protection (n = 7) |  |
| HEALTH FACILITY | OWNERSHIP | n | % | n | % | n | % |
| HCF-A | Private | 16 | 89 | 6 | 86 | 5 | 71 |
| HCF-B | Public | 18 | 100 | 6 | 86 | 5 | 71 |
| HCF-C | Public | 10 | 56 | 3 | 43 | 3 | 43 |
| HCF-D | Private | 11 | 61 | 6 | 86 | 4 | 57 |
| HCF-E | Private | 14 | 78 | 4 | 57 | 5 | 71 |
| HCF-F | Private | 16 | 89 | 6 | 86 | 6 | 86 |
| HCF-G | Private | 11 | 61 | 4 | 57 | 3 | 43 |
| HCF-H | Public | 14 | 78 | 3 | 43 | 4 | 57 |
| HCF-I | Private | 12 | 67 | 4 | 57 | 5 | 71 |
| HCF-J | Private | 12 | 67 | 5 | 71 | 5 | 71 |
| HCF-K | Private | 11 | 61 | 4 | 57 | 2 | 29 |
| HCF-L | Private | 11 | 61 | 4 | 57 | 3 | 43 |
| HCF-M | Private | 11 | 61 | 5 | 71 | 3 | 43 |
| HCF-N | Private | 13 | 72 | 5 | 71 | 4 | 57 |
| HCF-O | Private | 15 | 83 | 5 | 71 | 5 | 71 |
| HCF-P | Public | 14 | 78 | 5 | 71 | 6 | 86 |
| HCF-Q | Private | 11 | 61 | 4 | 57 | 3 | 43 |
| HCF-R | Public | 15 | 83 | 4 | 57 | 4 | 57 |
| HCF-S | Public | 14 | 78 | 4 | 57 | 4 | 57 |
| HCF-T | Private | 9 | 50 | 4 | 57 | 3 | 43 |
| HCF-U | Private | 12 | 67 | 4 | 57 | 3 | 43 |
| HCF-V | Private | 14 | 78 | 5 | 71 | 0 | 0 |
| HCF-W | Public | 15 | 83 | 3 | 43 | 4 | 57 |
| HCF-X | Private | 15 | 83 | 3 | 43 | 5 | 71 |
| HCF-Y | Private | 15 | 83 | 7 | 100 | 4 | 57 |
| **Public & Private** |  | **13** | **73** | **5** | **65** | **4** | **56** |
| **Public** |  | **6** | **31** | **4** | **57** | **4** | **61** |
| **Private** |  | **13** | **71** | **5** | **67** | **4** | **54** |

**Table 2:** Reported data on implementation of TB Infection Control measures in 25 health facilities stratified according to level of ownership

| Type of TB Infection Control measure assessed | Health Facility implementing IC measure (No, %) |  |  |  |  |  |
| --- | --- | --- | --- | --- | --- | --- |
|  | Public (n = 07) |  | Private (n= 18) |  | Total (n = 25) |  |
|  | YES | % | YES | % | YES | % |
| **Administrative/Managerial TBIC measures** |  |  |  |  |  |  |
| TB Infection Control (TBIC) Guideline available | 7 | 100 | 9 | 50 | **16** | **64** |
| TB Infection Control Committee available | 6 | 86 | 15 | 83 | **21** | **84** |
| TB Infection Control Plan available | 6 | 86 | 9 | 50 | **15** | **60** |
| TBIC plan displayed in a public place | 1 | 14 | 2 | 11 | **3** | **12** |
| TB Infection Control measures assessed within the last 6 months | 6 | 86 | 14 | 78 | **20** | **80** |
| Maintenance activities undertaken on structures so as to improve on TB infection control | 4 | 57 | 9 | 50 | **13** | **52** |
| Staff trained in TB infection control | 7 | 100 | 18 | 100 | **25** | **100** |
| All patients including newly diagnosed HIV+ clients screened for TB symptoms | 7 | 100 | 18 | 100 | **25** | **100** |
| Health education and/or counseling talks about TB infection control provided to TB patients | 7 | 100 | 18 | 100 | **25** | **100** |
| Health education materials on coughing etiquettes and respiratory hygiene available and strategically located at the health facility | 7 | 100 | 17 | 94 | **24** | **96** |
| Coughing patients provided with masks/tissues to reduce infectious aerosols | 7 | 100 | 13 | 72 | **20** | **80** |
| TB suspects/patients separated from other patients | 6 | 86 | 17 | 94 | **23** | **92** |
| MDR/XDR TB suspects isolated from other patients, including other TB patients and referred immediately for treatment | 2 | 29 | 6 | 33 | 8 | 32 |
| TB suspects prioritized for services to ensure shorter waiting time at the health care facility | 6 | 86 | 17 | 94 | 23 | 92 |
| Staff always reminded of the need for 'Early TB diagnosis' and 'Early TB treatment' | 7 | 100 | 17 | 94 | 24 | 96 |
| Properly ventilated and designated area for sputum collection available | 2 | 29 | 1 | 6 | 3 | 12 |
| A 'fast-queue' for collection of sputum smear results available | 5 | 71 | 13 | 72 | 18 | 72 |
| Sputum smear microscopy results reported within Turn Around Time (TAT) of 2 days | 7 | 100 | 16 | 89 | 23 | 92 |
| **Environmental Strategies to remove infectious aerosols after generation** |  |  |  |  |  |  |
| Windows in the consultation room, laboratory and waiting area able to open | 7 | 100 | 18 | 100 | 25 | 100 |
| Windows in the consultation room, laboratory and waiting area kept open during working hours | 7 | 100 | 18 | 100 | 25 | 100 |
| Fans used to increase circulation of air particularly in the consultation and waiting areas | 1 | 14 | 10 | 56 | 11 | 44 |
| Biosafety cabinet fitted with UV light available in the laboratory | 3 | 43 | 3 | 17 | 6 | 24 |
| Waste segregation system available | 7 | 100 | 18 | 100 | 25 | 100 |
| Infectious wastes treated before they are disposed off or incinerated/burnt | 3 | 43 | 14 | 78 | 17 | 68 |
| Open burning pit available | 0 | 0 | 5 | 28 | 5 | 20 |
| **Personal risk reduction strategies** |  |  |  |  |  |  |
| Staff often screened for TB infection symptoms | 3 | 43 | 6 | 33 | 9 | 36 |
| Staff often reminded or encouraged to know their HIV status | 6 | 86 | 16 | 89 | 22 | 88 |
| Staff often reminded of the risks of TB for people who are living with HIV | 6 | 86 | 16 | 89 | 22 | 88 |
| Staff often refresher trained on how to recognize and diagnose TB | 7 | 100 | 17 | 94 | 24 | 96 |
| N95 respirator/masks available at the point of assessment | 6 | 86 | 9 | 50 | 15 | 60 |
| Health care workers wear N95 respirators or masks | 3 | 43 | 5 | 28 | 8 | 32 |
| Staff fit tested on N95 respirators | 0 | 0 | 0 | 0 | 0 | 0 |

**Table 3:**
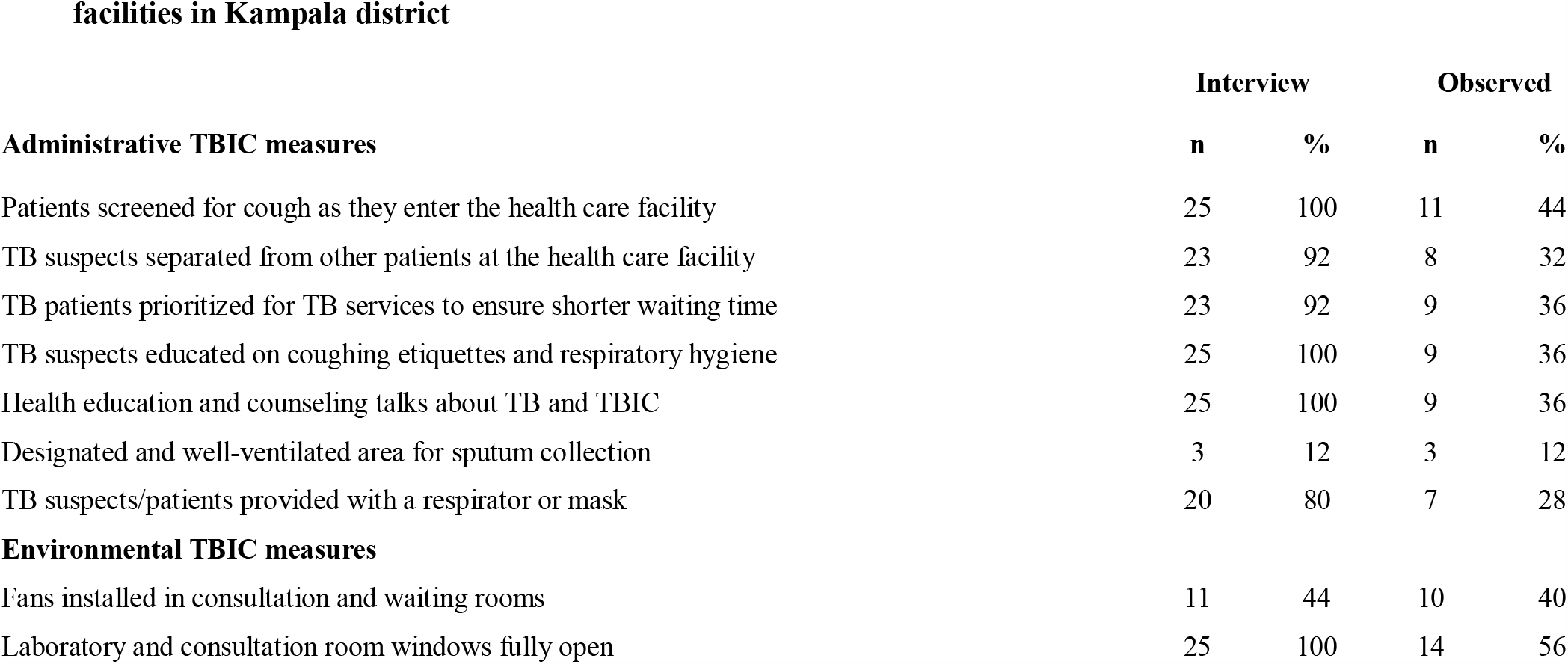
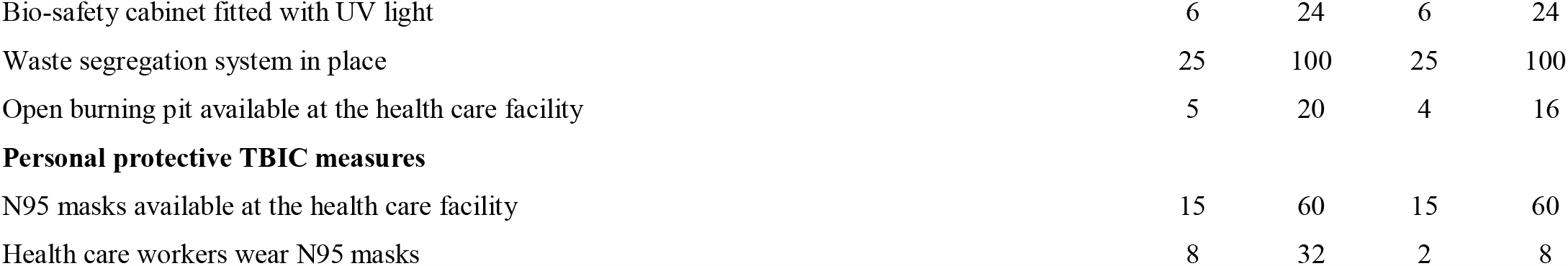
Reported and observed data on implementation of TB Infection Control measures in 25 health facilities in Kampala district

## Implementation of TB IC Measures

### Managerial TB Infection Control measures

#### Availability of TB IC guideline, committee and plan

Reportedly, 64% (16/25) of the health care facilities had a TBIC guideline, 84% (21/25) had a TBIC committee and 60% (15/25) had a TBIC plan at the time of assessment. A critical review of health care facility records revealed that, only 36% (9/25) of the Health facilities assessed had a TBIC plan and only 28% (7/25) had a TBIC guideline availed to the researcher. Of the 28% that evidently had a TBIC guideline, only 57% (4/7) had a national TBIC guideline of 2011 and the remaining 43% (3/7) had KCCA guidelines on TB infection control. Furthermore, only 12% (3/25) of the TB diagnostics and treatment health facilities assessed had a TBIC plan pinned on their notice boards or in a public place.

#### Health Care Facility TBIC Assessment and Infrastructure Maintenance Activities

Although 80% (20/25) of the Health facilities assessed reported that they had carried out a TB IC assessment on their health care facility infrastructures during the year, only one DTU presented evidence of the assessment and action plans were being implemented. The rest of the Health facilities stated that the assessment was carried out by NTLP/MOH officials and TB implementing partners, track TB in particular, but no findings had been communicated to them yet as evidenced by absence of TBIC assessment reports at the health care facility. Only 52% (13/25) of the health care facilities reported an improvement in infrastructure such as; acquisition of water proof tents for separation of suspected TB clients and confirmed TB patients (54%), acquisition of new fans so as to enhance sufficient ventilation in consultation rooms and waiting areas (30%), relocating laboratory room to a more spacious and properly ventilated area (8%), and lastly creating more windows on the TB/HIV clinic (8%).

### TB IC Trainings and Regular Screening of Health Care Workers for TB

From the survey findings, at least one health care worker from 96% (24/25) of the health care facilities assessed had received some level of training on TB IC from Ministry of Health and Regional Implementing partners. However, most Health facilities were not regularly screening for TB among health care workers. A one off TB screening among health care workers was reported in 36% (9/25) of the Health facilities assessed, usually at recruitment stage.

### Administrative TB Infection Control measures

#### Screening, separation and prioritization of Presumptive TB patients

The survey findings indicated that all Health facilities screened for TB signs and symptoms among all coughers at all entry points, 92% (23/25) separated presumptive TB patients from other patients and prioritized them first for services ahead of other category of patients so as to minimize their waiting time at the health care facility. By observation, 44% (11/25) were screening for TB signs and symptoms among patients, 32% (8/25) separated suspected TB patients from other patients and only 36% (9/25) prioritized TB patients (i.e. both TB suspects and those confirmed to be positive for TB) for TB services as soon as they arrived at the health care facility. It was further observed that some TB patients were sitting in the same waiting area (water proof tent) together with other vulnerable patients (HIV positive clients). This malpractice was particularly observed at high volume government and faith based-private not for profit health care facilities.

#### Health Education and Counseling on respiratory hygiene and coughing etiquettes

Although all Health facilities reported that they were providing health education and Counseling to confirmed TB clients and presumptive TB patients, Health workers were observed giving health Education messages on coughing etiquettes and respiratory hygiene to this category of patients at 36% (9/25) of the Health facilities assessed. However, it is imperative to note that some Health facilities did not have presumptive TB patients or confirmed TB patients on the day of health care facility visit. More specifically, private health facilities reported that they rarely received TB patients or suspects on a daily basis. These further made it difficult to observe whether health education and counseling talks were provided at these health care facilities. In addition, some health facilities were assessed in the morning and evening hours and on certain occasions during weekend days, thus contributing to the low level of observed TBIC practices in some TB centers assessed.

#### Availability of Information, Education and Communication (IEC) materials

Ninety six percent (24/25) of the health care facilities assessed had IEC/BCC materials (i.e. flip charts, TB screening patients’ flow charts, desk booklets and TB fact sheets) containing messages on TB Infection Control. Worth mentioning, TB IEC materials were found kept in the drawer at the reception and others in the consultation rooms in two private health facilities. In addition, some health care facilities had received DVDs containing TB messages (Both in Luganda and English languages) and TV sets from USAID funded Track TB project, but unfortunately, none of them was found displaying TB messages at the time of assessment. Similarly, in some private health care facilities with audio-visual aids, patients were often observed watching other local TV channels in the waiting area.

#### Early Diagnosis and Prompt Treatment

Early diagnosis and prompt treatment forms the first pillar of the End TB strategy. Most health facilities (92%) assessed disseminated sputum smear microscopy results within 24 hours from the time of collection of sputum specimen, and whenever a patient was found positive, treatment was initiated on spot or within not more than 24 hours.

#### Availability of a well Ventilated and designated area for Sputum Collection

A few health care facilities (12%, 3/25) surveyed had a designated area for collection of sputum specimens. Health facilities that didn’t have designated areas for sputum collection often instructed patients to collect sputum from under the tree or behind the health care facility premises. One private health care facility, often instructed presumptive TB patients to collect sputum specimen from the toilet possibly due to stigma.

### Environmental TB Infection Control measures

#### Natural Ventilation (Window Opening)

Although all health facilities reported that they open their windows during working hours, only 56% (14/25) had windows fully opened on the day of assessment. At one health care facility, windows were fitted with wire mesh which interrupts about 30% of the natural air to move freely in and out of the room. Sliding windows were also observed in some health facilities, moreover with the middle part of the window completely fixed. At the outpatient department, most Health facilities had properly ventilated waiting areas particularly for presumptive TB cases. However, it is imperative to note that some of these waiting areas were improvised water proof tents supplied by Track TB, USAID and MSH and others ordinary tents improvised by the health care facilities themselves.

#### Availability of Bio-safety cabinets (BSC)

A few Health facilities (24%, 6/25) had bio-safety cabinets fitted with ultra-violet (UV) light and often handled sputum specimens inside these safety Cabinets. BSC users reported that they often switched on the UV light overnight in order to disinfect the work surfaces inside the bio-safety cabinets. However, there was no evidence to show that these bio-safety cabinets were regularly serviced by certified servicing companies. Further, one laboratory technician was observed working in a bio-safety cabinet which was cluttered with sputum mugs, urine containers, boxes of gloves and other un necessary materials. This can interrupt the air follow inside the bio-safety cabinet eventually letting some contaminated air to come outside the bio-safety cabinet.

#### Mechanical Ventilation Equipment availability

By observation, only 40% (10/25) of the health care facilities had fans installed either in the waiting area, consultation rooms or both. Though at some health care facilities assessed, it was reported that installation of fans in the waiting areas and consultation rooms was not necessary because they could increase one’s risk of contracting TB disease. The availability of HEPA filters was not assessed since practically very many facilities cannot afford to install them in their places of work due to the high costs involved in purchasing them.

#### Waste Management practices

Most health care facilities assessed had commendable waste segregation practices. Every DTU had a sharps container, a non-infectious, less infectious and highly infectious waste collection containers. However, bin liners with appropriate color coding were lacking at some Health facilities assessed, making it difficult for support staff (cleaners) to distinguish biohazard from non-biohazard wastes during the cleaning process. During data collection period, 68% (17/25) disinfected Sputum specimens before disposal with varying concentrations of JIK. Burning pits were observed at 16% (4/25) of the Health facilities assessed, 8% (2/25) had an autoclave, 68% (19/25) had contracted a biohazard waste collection company that collected infectious wastes on a bi-weekly basis, 8% (2/25) neither had a burning pit, an autoclave nor a contract with biohazard waste collection company. In the later, one DTU was observed dumping sputum sediments or remains into the placenta pit, the other facility was simply burning medical wastes including sputum containers on a leveled surface within the health facility premises.

### Personal protective measures

According to the survey findings, 60% (15/25) of TB diagnostics and treatment health care facilities had N95 respirators in place. Of these, only 53% (8/15) reported that their health care workers wore N95 respirators when talking to presumptive or confirmed TB clients or even when working on the TB sputum specimens in the laboratory.

By observation, respirators were worn by health workers at only 13% (2/15) of the health facilities with adequate stocks of respirators. In addition, no respirator fit testing was conducted at any of the health care facilities assessed, which further increase one’s chances of contracting TB infection. At one of the government owned facilities, we observed some patients putting on masks while others not, in the same waiting area. More so, some patients were repeatedly observed sliding their masks just below their mouth whenever they were coughing, others walked around while holding masks in their hands, where as some female patients deliberately kept their masks in their hand bags and only worn them whenever a health worker visited the tent they were sitting. One patient deliberately refused to put on a mask when she was told to do so and simply walked away with it in the hands as she was going to expectorate the sputum specimen. Lastly, a laboratory personnel was found wrapping a handkerchief around the facial area instead of N95 respirator to protect himself from TB Infection.

## Discussion

This study is the first of its kind to document the extent of implementation of IC guidelines in both public and private health care settings in Uganda. Our findings indicates that administrative/managerial (73%) and environmental (65%) were the most Implemented IC measures at the health facilities assessed. Personal protective TB IC measures (56%) were the least implemented measures, consistent with other studies in low resource, high TB prevalence settings [18, 19]. Further, no health care facility implemented all the three levels of TB infection control (i.e. administrative, Environmental and Personal protective measures) in an integrated manner. Gaps in implementation of TBIC measures in TB health care settings predisposes health professionals and other patients to nosocomial tuberculosis transmission [20].

Although administrative TB IC measures were the most implemented, study findings revealed that only 31% of the recommended administrative measures were being implemented at public health care facilities compared to 71% in private health facilities. This finding can possibly be attributed to the fact that public health facilities were overcrowded with patients probably due to inadequate Human resource, thus, limiting the implementation and close monitoring of certain administrative measures like triaging and respiratory hygiene respectively. The lack of properly ventilated and designated area for sputum collection further curtails the TB infection control efforts at both public and private health facilities

In regards to screening of health workers policy, the Uganda national TB Infection Control guidelines (2011) recommends regular screening of health workers for TB disease using intensified case finding guide (ICF) symptom screening tool on a five year basis or when indicated and on every new staff. However, this was not being implemented at all at 64% of the health facilities assessed. This finding is not very uncommon because existing literature suggest that most health workers often do not feel comfortable to access TB and HIV screening, diagnosis and treatment services at their health facilities of employment [21]. In this regard, compulsory routine screening for TB among all health workers need to be institutionalized and monitored by the National TB Program

Our findings suggest suboptimal implementation of environmental IC measures. Only 56% of the health care facilities routinely opened their windows while 40% had improved their ventilation mechanically by use of fans. The use of poorly designed or poorly maintained ventilation systems, leading to inadequate airflow, can result in health care associated transmission of M. tuberculosis.

Notable discrepancies in observed data and survey data were documented. This illustrates that HCWs relatively know what they are expected to do in terms of TB IC, but probably because of failure to appreciate the importance of TB IC, recommended measures are not implemented. This discrepancy between reported and observed measures also highlights the weakness of self-reports [22].

In addition, several research studies [18, 23, 24] posits that some health workers who are knowledgeable about TB infection control practices do not implement them at their places of work. This highlights the need to explore other barriers and facilitators for TB IC implementation at TB Diagnostic and Treatment health facilities in Ugandan context through use of qualitative approaches so as to inform development of targeted strategies aimed at curtailing the high risk of transmission of TB Infection within Diagnostics and treatment units.

There are several important limitations of this study. Most measurements relied on self-reporting, introducing the possibility of social desirability bias. We attempted to minimize this bias by including objective observations in our analysis. In addition, ventilation adequacy was only determined by presence of windows and ventilators in the waiting areas, consultation and laboratory rooms. However, the researcher did not assess the appropriateness of air ventilation using the ratio of window area to the floor/room area as stipulated in the Uganda’s TB infection control guidelines (2011).

## Conclusion

Generally, implementation of WHO recommended TB IC measures in health facilities delivering TB care services in Kampala was sub optimal. Routine involvement of health facility in-charges, managers and administrators as well as increasing human resources for health is critical in implementing easy to do TBIC measures like triaging, patients’ educating on coughing etiquette and respiratory hygiene and daily window opening particularly in public health care settings where implementation of administrative TB IC measures is wanting. Our study findings suggests participatory in-service training for health workers focusing on implementation of infection control practices and where feasible, install upper-room germicidal ultraviolet (GUV) systems most especially in public health care settings with overcrowding and inadequate ventilation. Lastly, we recommend further explorative studies aimed at investigating the associated determinants responsible for the observed level of implementation of TB IC measures in a similar setting.

## Data Availability

The data that support the finding of this study is available from the corresponding author and can be provided upon reasonable request

## Declarations

### Ethics approval and consent to participate

Ethical review and approval was obtained from The Uganda Martyrs University Faculty of Health Sciences Higher Degrees Research and Ethics Committee. In addition, permission to conduct this study was sought from the Directorate of Public Health and Environment, Kampala City Council Authority (KCCA), the governing body for Kampala city. Furthermore, informed written and verbal consent was obtained from the participants at the time of data collection. Confidentiality and use of data for research purposes was further emphasized prior to the beginning of the data collection.

### Consent for publication

Not applicable.

### Copyright

The Corresponding Author has the right to grant on behalf of all authors and does grant on behalf of all authors, a non-exclusive license on a worldwide basis to the BMJ Publishing Group Ltd to permit this article (if accepted) to be published in BMJ editions and any other BMJPGL products and sublicenses such use and exploit all subsidiary rights

### Author’s contributions

**DT** took lead in conceptualization of the study idea, protocol write up, data collection, compilation and conception of the manuscript idea and its write up. **SPK** Supervised and guided the protocol write up, data collection, compilation and conception of the manuscript idea and its write up. **LB** and **MK** participated in the review and development of the study proposal, reviewed and guided the compilation and conception of the manuscript idea and its write up. **ST, SZM, RKM, SM, AN, JK, DM** contributed to the drafting and subsequent revision of the manuscript. All authors read and approved the final manuscript

## Acknowledgement

The authors would like to thank all the study participants/respondents for fully accepting to participate in this study, Uganda Martyrs University Research institutional Review Board and KCCA directorate of public health and environment whom we sought permission to conduct this study.

